# Dopamine D_1_ agonist effects in late-stage Parkinson’s disease

**DOI:** 10.1101/2022.04.30.22270885

**Authors:** Mechelle M. Lewis, Lauren Jodi Van Scoy, Richard B. Mailman, Sol De Jesus, Jonathan Hakun, Lan Kong, Yang Yang, Bethany Snyder, Sridhar Duvvuri, David L. Gray, Xuemei Huang

**Author notes:** These authors contributed equally to this study (Mechelle M. Lewis, Lauren Jodi Van Scoy, Richard B. Mailman). Correspondence to the following authors: Mechelle M. Lewis, Ph.D., Penn State Hershey Medical Center, 500 University Dr., H-037, Hershey, PA 17033-0850, Xuemei Huang M.D., Ph.D., Penn State Hershey Medical Center, 500 University Dr., H-037, Hershey, PA 17033-0850.

## Abstract

**Background:** Current pharmacotherapy has limited efficacy and/or intolerable side effects in late-stage Parkinson’s disease (LsPD) patients whose daily life depends primarily on caregivers and palliative care. Clinical metrics inadequately gauge efficacy in LsPD patients.

**Objective:** Explore if a D_1/5_ dopamine agonist will have efficacy in LsPD that will be detected most sensitively by caregivers in a phase I study.

**Methods:** A double-blind controlled phase Ia/b study compared the D_1/5_ agonist PF-06412562 to levodopa/carbidopa in six LsPD patients. Throughout the study, caregivers were with the patients. Assessments included standard quantitative scales of motor function (MDS-UPDRS-III), alertness (Glasgow Coma and Stanford Sleepiness Scales), and cognition (Severe Impairment and Frontal Assessment Batteries) at baseline (Day 1) and thrice daily during drug testing (Days 2 and 3). Clinicians and caregivers completed clinical impression of change questionnaires, and caregivers participated in a qualitative exit interview. Blinded triangulation of quantitative and qualitative data was used to integrate findings.

**Results:** Neither traditional scales, nor clinician impression of change, detected consistent differences between treatments in the five participants who completed the study. Conversely, the overall caregiver data strongly favored PF-06412562 over levodopa in four of five patients. The most meaningful improvements converged on motor, alertness, and engagement.

**Conclusion:** D_1/5_ agonists may offer potential benefit for LsPD patients. Caregiver perspectives with mixed method analyses may overcome limitations in standard rater/clinician-based evaluations. Further studies are warranted and need to integrate caregiver input as an essential component of outcome evaluations.

**Trial Registration#:** ClinicalTrials.gov: NCT03665454

## Introduction

Parkinson’s disease **(PD**) is characterized clinically by motor and non-motor symptoms. Despite research advances related to disease modifying therapy, symptomatic therapy using the dopamine precursor levodopa remains the cornerstone of therapy.^1^ Unfortunately, progressive dopamine neuron loss markedly decreases bioconversion of levodopa to dopamine in the striatum but not mesolimbic areas, thereby decreasing efficacy and increasing side effects.^2^ Additionally, potential “off-target” effects from dopamine formation in other monoamine neurons may cause side effects such as drowsiness and hallucinations.^3-6^ Late-stage PD (**LsPD**) patients also experience many non-motor symptoms including anxiety/depression, pain, sleep disorders, cognitive decline, apathy, and social withdrawal,^7,8^ some of which predate motor dysfunction.^9^

As PD patients advance to LsPD, there is a high burden for family and caregivers, and higher healthcare costs compared to early- and mid-stage (sometimes called advanced) patients.^10-15^ There have been no prior controlled trials of drugs in LsPD patients due in part to the perceived fragility of the patients, lack of validated assessment tools for LsPD, and no accepted target that might mediate symptomatic benefit in patients where levodopa has limited efficacy. Because post-synaptic cytoarchitecture in LsPD patients is largely preserved despite dopamine neuron degeneration, targeting one or more of the post-synaptic dopamine receptor populations theoretically could offer marked therapeutic benefit.

Although many “dopamine agonists” are approved clinically and have some utility in earlier-stages of PD, they have inferior efficacy to levodopa and intolerable side effects in LsPD.^16,19,20^ The term “dopamine agonist,” however, is misleading -- currently approved “dopamine agonists” are selective for dopamine D_2_-like receptors (e.g., see Supplemental Table A).^16-18^ There is compelling neurobiological and pharmacological evidence for the potential of D_1_ receptor-selective agonists in LsPD.^19,21,22^ This is supported by experimental data in severe MPTP-treated non-human primates (NHPs)^23,24^ and mid-stage PD patients.^25,26^ The classical experimental D_1_ agonists, however, contained a catechol-moiety that came with significant pharmaceutical liabilities.^19^ Newer D_1_ agonists have overcome this limitation^27^ and shown efficacy in early- or mid-stage PD patients.^28-31^ Thus, they also may have utility in LsPD patients.

Our search of the literature found no prior interventional studies conducted in LsPD. The accessibility of an orally available D_1/5_ partial agonist PF-06412562 (*henceforth PF-2562*) allowed us first to evaluate the safety of a D_1/5_ agonist in a very short (two-day) feasibility phase I pilot study of LsPD patients whose results were reported.^32^ We now explore the efficacy of PF-2562 from that study using several *a priori* postulates. The first was a primary focus on caregiver impressions^33-36^ because they were most familiar with patient behavior. In addition, standard clinical metrics obtained over the two-day study were less likely to capture meaningful changes in LsPD patients who had severe multi-domain disability. Finally, a convergent mixed methods design was used that involved collecting <u>both</u> quantitative and qualitative data, and comparing and contrasting findings.^37^

## Patients and Methods

### Study design, subjects, and randomization

This study was conducted at PennStateHealth (PSH) in compliance with the ethical principles of the Declaration of Helsinki and guidelines for Good Clinical Practice issued by the International Conference on Harmonization. It was reviewed and approved by the US Food and Drug Administration and PSH Institutional Review Board. All participants and/or their caregivers provided signed informed consent. Details of subject recruitment, inclusion and exclusion criteria, baseline medical, protocol information, and safety data were published in a previous report.^32^ Briefly, all participants were recruited from our Movement Disorders clinic or via a local PD support group and met published diagnostic criteria.

All LsPD subjects had disease duration >15 y and Hoehn & Yahr stages ≥IV, either “on” or “off” levodopa. Our criteria were similar to those used by Coelho and Ferreira,^38^ but differed from others who have used this term less specifically (e.g., disease duration <5 y and Hoehn & Yahr stages II-III^39^). As a condition of their participation, all subjects were informed that regardless of response to PF-2562, they would not be permitted to continue taking PF-2562. After informed consent, participants and caregivers were admitted to the Clinical Research Center (CRC) for four days during two consecutive weeks. To maximize comfort, levodopa/carbidopa (for parkinsonian symptoms), acetaminophen (for pain), ondansetron (for nausea), and diphenhydramine (for allergies) were given throughout the study when needed.

Eligible participants were randomized to either PF-2562 (Sequence A) or levodopa (Sequence B) during the first test period using a 1:1 random allocation sequence, and then crossed-over to the other drug during the second test period (see Supplemental Figure A). Participants, caregivers, and investigators were blinded to sequence assignment, and participants received identical numbers of pills (i.e., containing PF-2562, levodopa, and/or placebo) that were administered at the same time during each sequence. Specifically, following overnight levodopa/dopamine agonist washout and baseline evaluation on Day 1, participants assigned Sequence A received PF-2562 (25 mg at ∼0900 h and 20 mg 4 h later) on Days 2-3 during Test Period 1, whereas they received encapsulated Sinemet (carbidopa/levodopa 25/100 mg) 3-4 times (depending on pretrial regimen) 4 h apart on Days 2-3 during Test Period 2. Participants assigned Sequence B received Sinemet in Test Period 1 and PF-2562 in Test Period 2. On Day 4, all participants resumed pre-trial treatment and were discharged after demonstrating no significant complications.

### Choice of study compound

The initial pilot study focused on establishing the safety and tolerability of a D_1_ agonist in LsPD, thereby querying the feasibility of conducting clinical trials in LsPD. Among the available D_1_ agonists, PF-2562 was selected because it caused acute antiparkinsonian effects in 13 PD patients and was well-tolerated at a 50 mg oral dose (t_½_=6.4 h) split into 30 and 20 mg doses given four hours apart.^29^ This informed the current study design and split-dose regimen involving a short in-patient stay and cross-over design. Tavapadon, a related D_1_ agonist, is dosed via a titration regimen to reach efficacious drug levels,^31^ and the limitations of this pilot study did not allow for extended in-clinic stays to accommodate titration.

### Quantitative data and metrics

We included five standard quantitative scales^40-43^ for specific efficacy domains representing: motor [MDS-UPDRS motor scale (MDS-UPDRS-III)]; alertness [Glasgow Coma (GCS) and Stanford Sleepiness (SSS) Scales]; and cognition [(Severe Impairment (SIB), and Frontal Assessment (FAB) Batteries]. Scores were obtained three times each on Days 2-3: prior to drug administration and then one hour after the first and second dose. We also evaluated sleep using polysomnography (PSG), except in two participants (3 and 4) with deep brain stimulation due to disruption of EEG signals captured during PSG. From these data, “sleep efficiency” was selected as the most global/comprehensive metric to report.

As detailed in our previous report,^32^ movement disorder clinicians and caregivers completed an adapted validated global clinical impression (GCI) scale designed to assess severity (GCI-S) or change (GCI-C). <u>On Day 1</u>, clinicians evaluated patients’ history and exam (H&P), and summarized it as a single GCI-S score ranging from 1=normal/not ill to 7=extremely ill. Caregivers completed a baseline GCI-S on Day 1 based on their knowledge of the participant’s disease at home that included 17 items we summarized as one score ranging from 0-102. <u>At the end of Days 2-3,</u> both caregivers and clinicians completed the GCI-C questionnaire that included 17 items scored on a 7-point Likert scale (−3=marked worsening; 0=no change; 3=marked improvement). Clinicians completed this assessment based on interviews with caregivers and daily examination of patients. As pre-specified, Day 3 metrics were used for final analyses to avoid likely confounders such as excitement/noise/environment changes on Day 2.

### Qualitative interviews

Qualitative data collection was chosen to capture broad, nuanced experiences, observations, and perspectives of caregivers regarding potential efficacy and/or side effects. Semi-structured caregiver interviews (30-60 minutes) were conducted by a trained qualitative research assistant at the end of Day 3. Responses were audio-recorded and transcribed verbatim. Interviews explored the extent to which caregivers perceived patient response to study drug (if any) and adverse effects compared to patient status at baseline. The interview guide used open-ended questions to elicit first general observations from caregivers and then probed specific domains of motor, alertness, cognition, and sleep (*vide infra*).

### Convergent mixed methods design

Convergent mixed methods designs collect both quantitative and qualitative data for a ‘domain’ of interest and then compares and contrasts the conclusions from each dataset (‘merging’) to reach a comprehensive conclusion.^44^ At study conception, pre-selected domains were guided by our clinical experiences with LsPD patients and extant literature. Table 1 lists these domains of interest (motor, alertness, cognition, sleep, and clinician/caregiver impression of change), and the quantitative and qualitative measures that corresponded to each domain. Domains were analyzed separately and conclusions drawn independently. Blinded data then were integrated by merging findings and seeking points of convergence or divergence in the conclusions.^44,45^ This mixed methods approach establishes stronger credibility and validity to the findings when convergence of conclusions is established and opportunities to extract lessons when divergence is detected.^44,45^

**Table 1.**
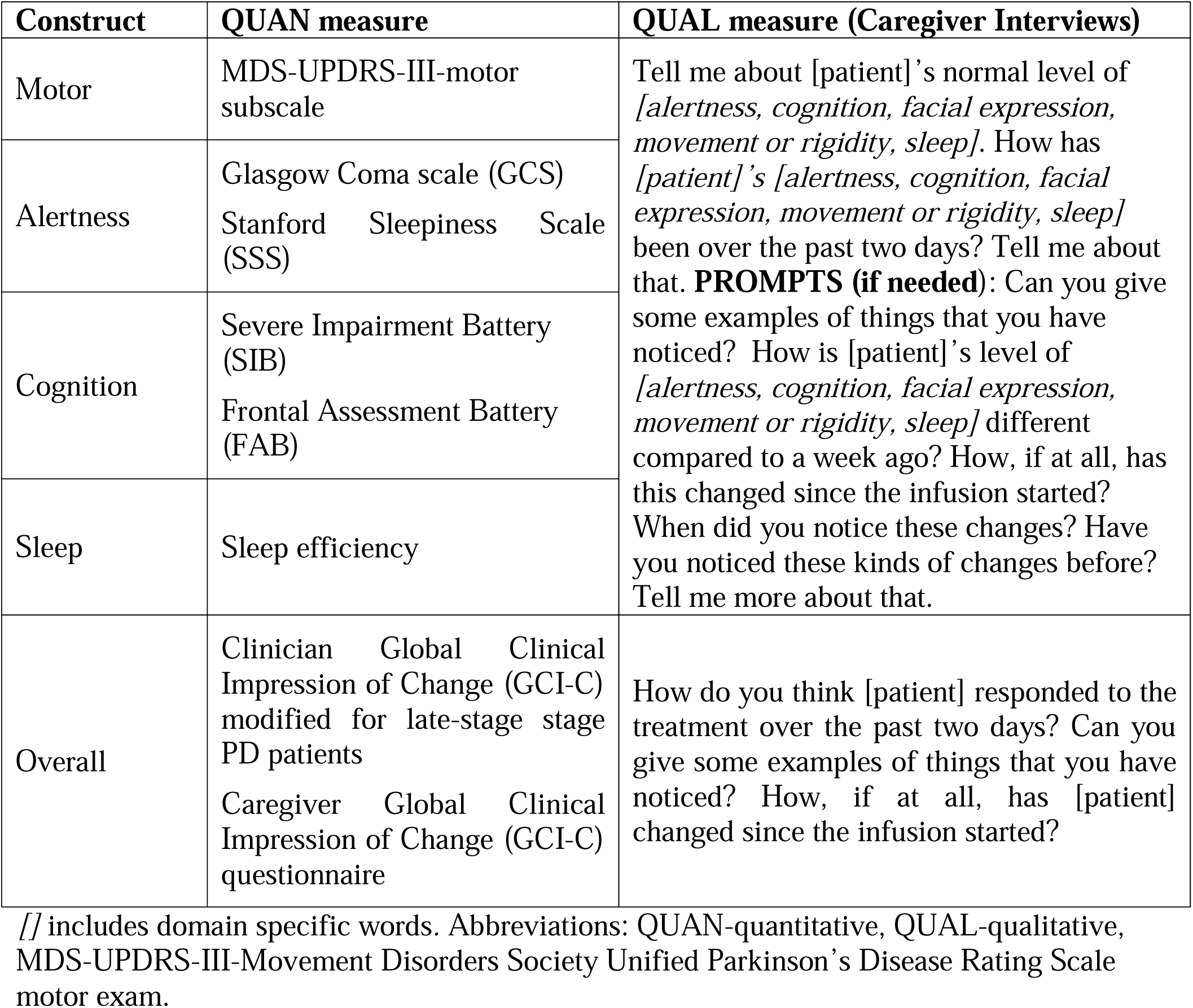
Convergent mixed methods study design: Constructs and Measures.

### Analysis

#### Quantitative analysis

Quantitative scales provided either one score (GCS, SSS, and sleep efficiency) or several that were summed (SIB, FAB, and MDS-UPDRS-III) for each participant. The score at the start of Day 2 prior to study drug administration then was subtracted from the score at the end of Day 3 to evaluate change. Both clinician and caregiver GCI-C scores also were captured. For the purposes of this study, we included motor, alertness, cognition, and sleep scores. Scores are presented for each participant in this pilot study of six subjects. Based on pre-determined efficacy assessment, the primary endpoint was caregiver ratings that were analyzed using a paired Student’s t-test (two-tailed α=0.05).

#### Qualitative analysis

Conventional content analysis including data transformation was used to evaluate the data.^44^ Published guidelines for methodological rigor of qualitative analysis were followed to ensure attention to the truth-value, applicability, consistency, and neutrality of findings.^44,46^ Three independent, blinded analysts used qualitative software (NVivo Ver. 11.0, QSR International, Melbourne Australia) to code and analyze the data. Analytic details are provided in Supplemental Text A.

#### Mixed methods integration

Joint displays were constructed to compare quantitative efficacy outcomes with results from the transformed qualitative data for each participant completing the study (n=5). The study team reviewed conclusions from both the quantitative and qualitative datasets to ascertain an integrated conclusion of the preliminary efficacy of PF-2562.^45^

### Data sharing

De-identified data supporting the findings of this study are available upon reasonable request from corresponding author XH. The data are not publicly available due to privacy or ethical restrictions. All requests must be in writing and will be evaluated in a timely manner by the TBRC executive team.

## Results

### Participants

Six subjects met inclusion criteria (demographics in Supplemental Table B). Patients had a mean age of 73.5 (±4.5 SD) y and two participants were female. Consistent with protocol inclusion criteria, patient Hoehn & Yahr stages all were >4 in the ‘on’ state. No subject required levodopa rescue during the PF-2562 week, whereas one participant was administered rescue medication during the levodopa week (subject 4, 0.5 100/25 mg levodopa/carbidopa tablet on Day 2, 1 on Day 3). Of the six patients who were randomized, one (subject 6, disease duration 19 y) withdrew after the first arm because of blood pressure fluctuations the clinical team felt were related to the interaction of test drug with baseline dehydration, related kidney dysfunction, and autonomic dysfunction.^32^ This patient’s data is excluded from these efficacy analyses. The remaining five patients completed both arms of the study.

Key narrative phrases from caregiver interviews qualitatively describe the patient’s baseline functional status (Supplemental Table B). Four of five patients (subjects 1, 3, 4, and 7) represented classic LsPD patients and had disease durations ranging from 15-23 y. All patients had been treated with symptomatic drugs and two with deep brain stimulation. In addition to motor disability and requirement of walker and/or wheelchair use, all patients had varying challenges in maintaining normal sleep/wake cycles, being alert and engaged during daytime, and all displayed clinically significant cognitive dysfunction.

Subject 8 had the longest disease duration (32 y). All drugs including levodopa had caused intolerable side effects and thus this patient had not been treated with any parkinsonian drugs for three years prior to study enrollment. On most days, he was in unarousable “deep sleep,” but was able to reflexively suck and swallow if his mouth was stimulated with a straw or food in a more “awake” state. Because of his atypical background and long survival without dopaminergic medication, we highlight his response to treatment in subsequent sections because it may provide unique insight into the pathophysiology of LsPD.

### Quantitative results

Standardized scales assessing motor function, alertness, cognition, and sleep did not detect a clear pattern of differences between levodopa and PF-2562 (see details in Supplemental Text A and Table 2). Clinician scores were more variable than those from caregivers for both levodopa and PF-2562 (Figure 1A), and caregivers rated PF-2562 consistently better than levodopa (p=0.007; Figure 1B). This offered initial evidence that PF-2562 may provide improved efficacy based on caregiver scores.^32^

**Table 2:**
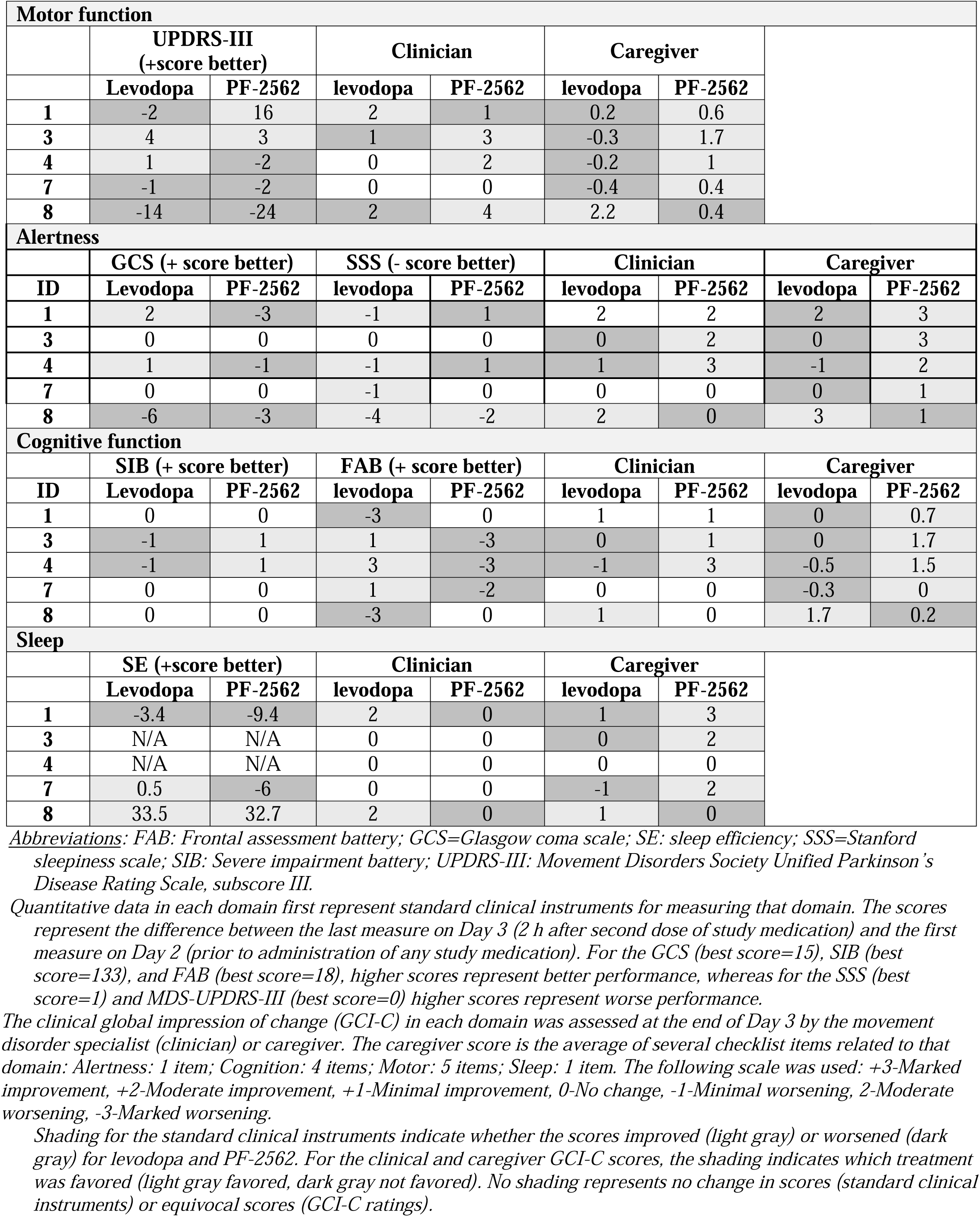
Summary of quantitative data.

**Figure 1.**
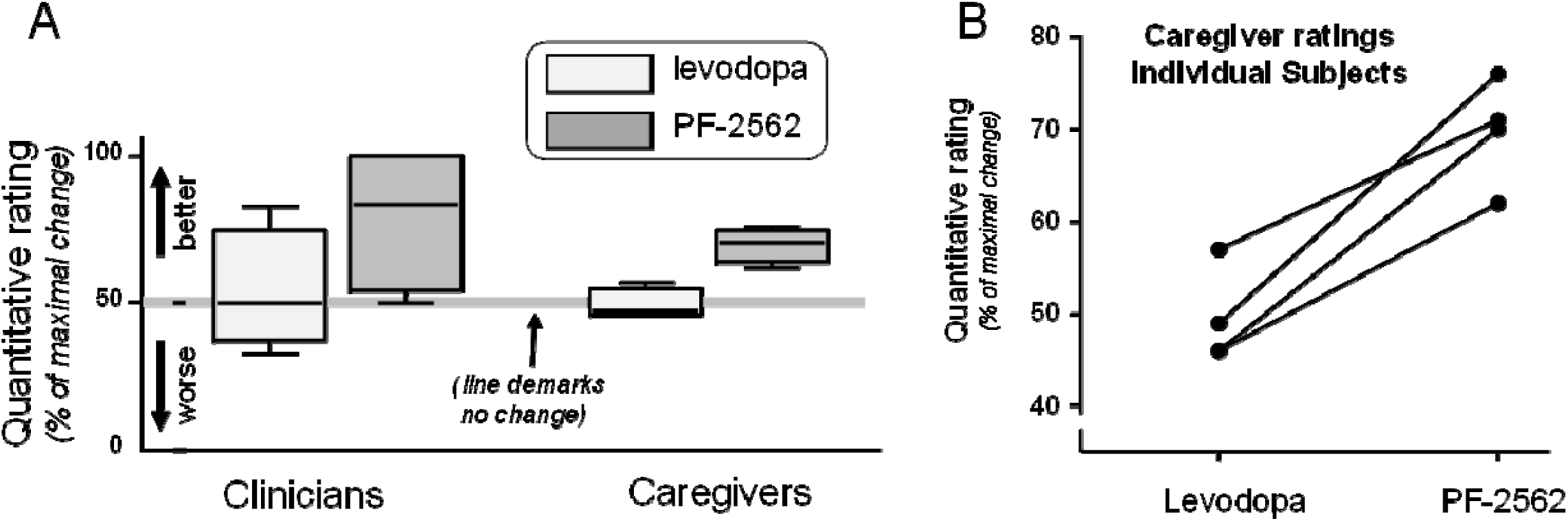
Evaluation of quantitative GCI data for four of five subjects.

### Qualitative caregiver interview results

Blinded analysis of the transcripts revealed significant variability in patients’ baseline functional status (see details in Supplemental Text B and Table 3). Notably, caregivers did not distinguish explicitly among alertness, attention, and cognition according to qualitative analyses. Therefore, these domains were collapsed as ‘patient overall engagement’ in the mixed methods joint display. Results of the qualitative data transformations (improved, worsened, unchanged) are shown in Table 3, along with quotations from caregivers describing the changes they noticed within each domain. Overall, the qualitative data suggested that PF-2562 improved cognitive engagement and motor domain status (balance, weakness, and rigidity) in the four typical LsPD subjects.

**Table 3.**
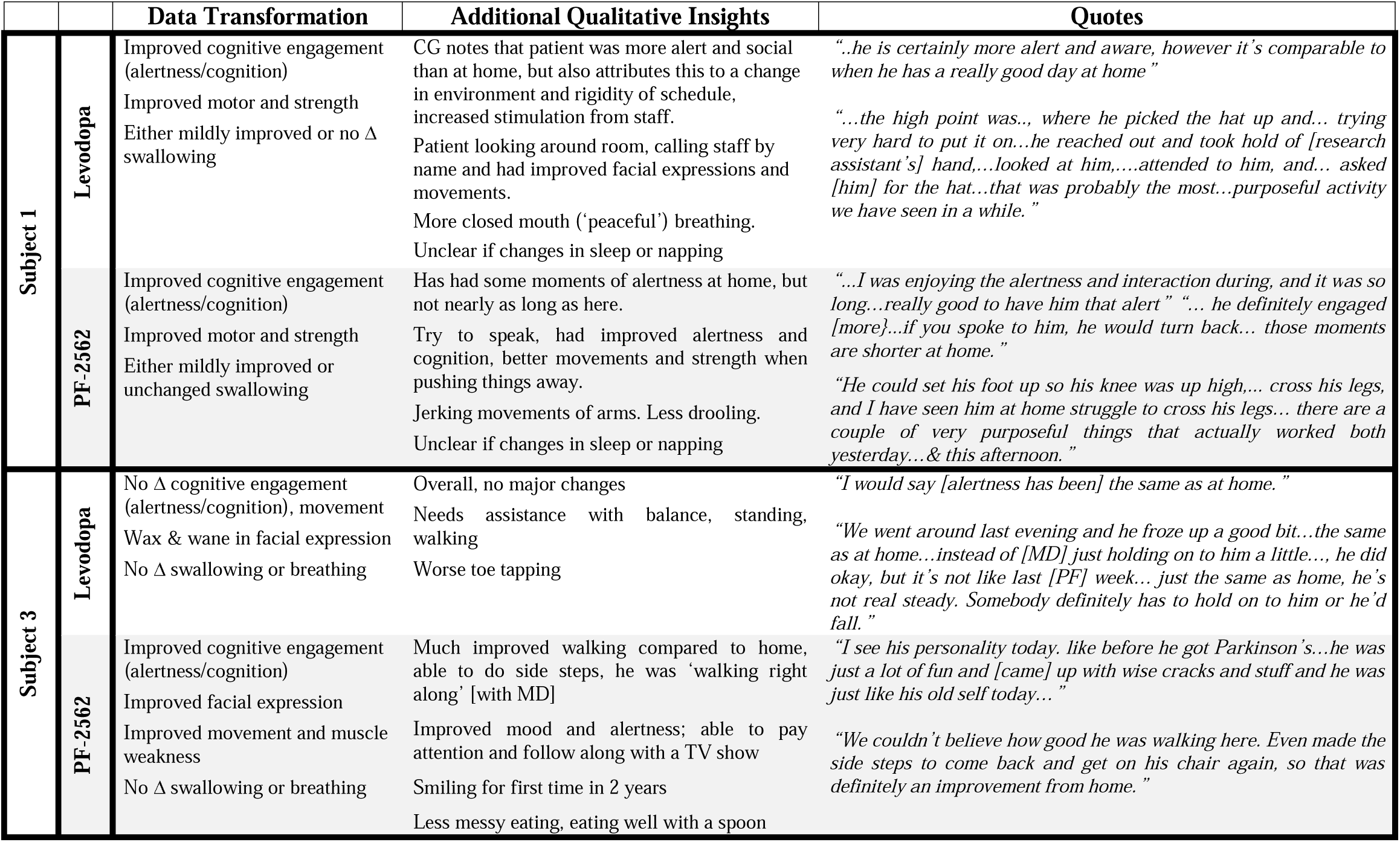
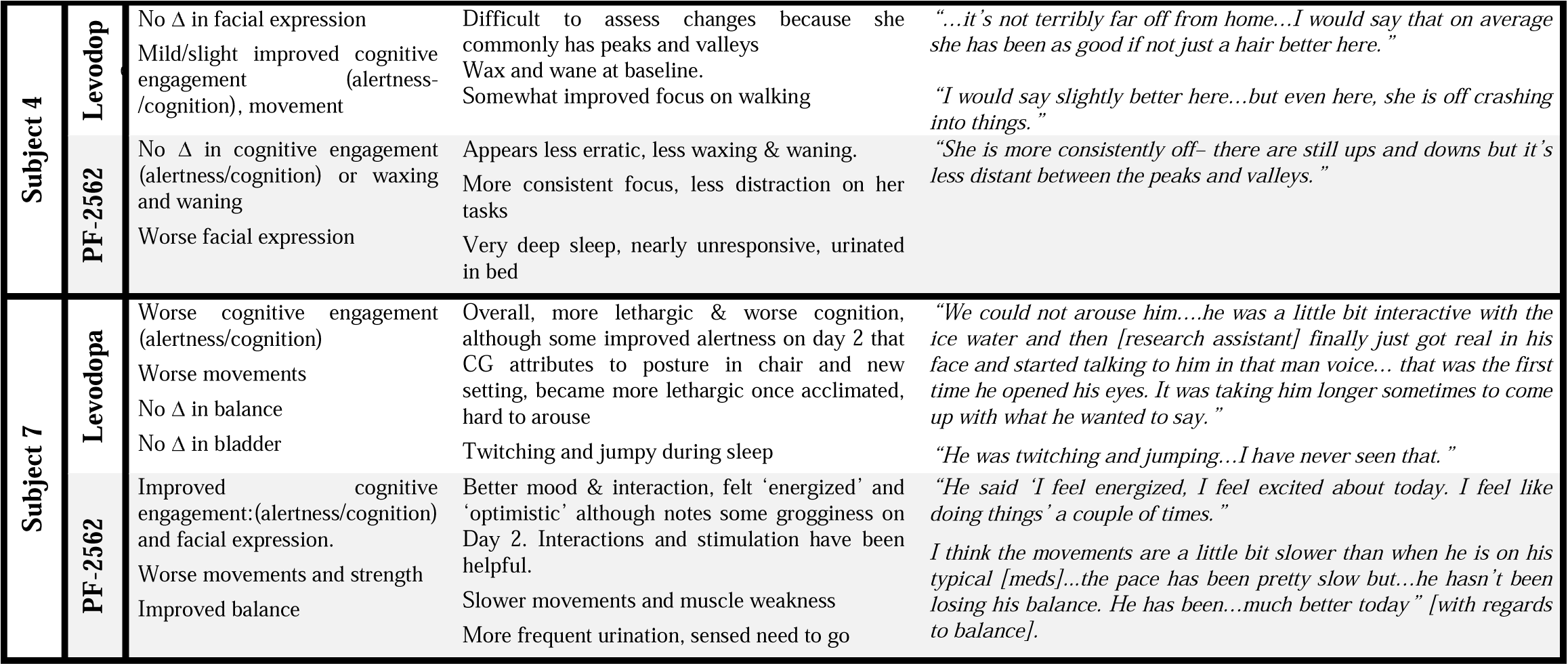
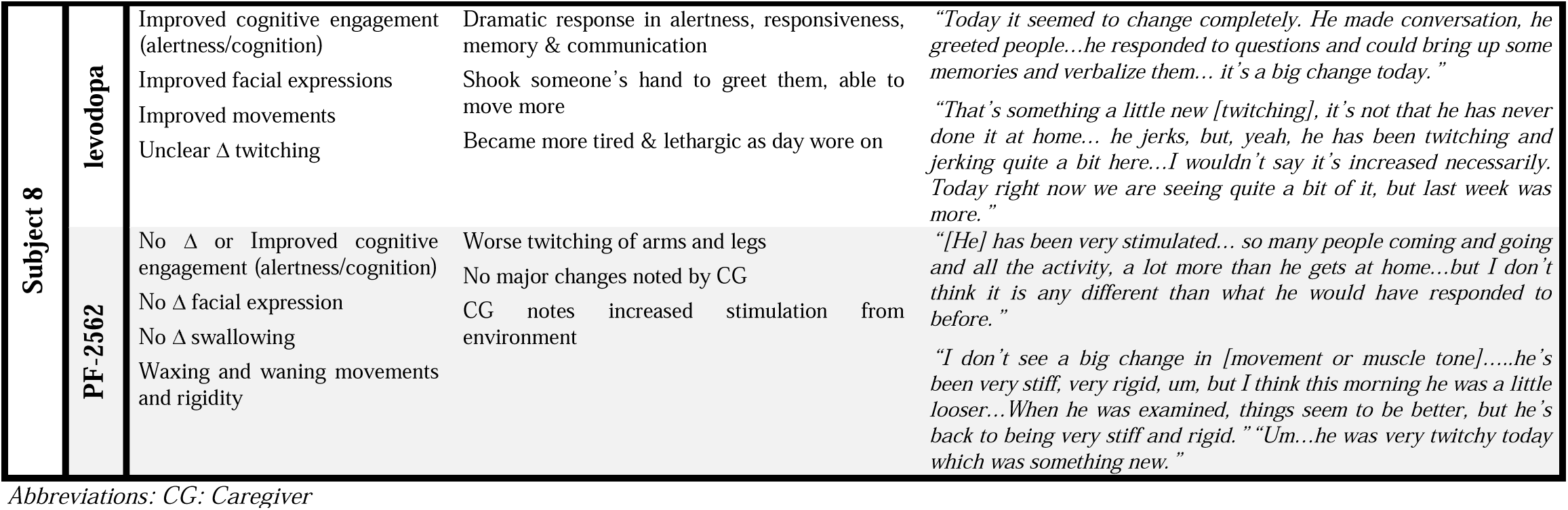
Qualitative data transformation and quotations (PF-2562 in grey cells)

Qualitative analyses also suggested that PF-2562 may improve facial expression and sleep to varying degrees, although qualitative analysis of sleep was challenging due to highly variable caregiver descriptions (e.g., judgment of sleep quality based on different aspects such as breathing, apneas, duration, depth of napping, restlessness, vocalizations). All caregivers commented that some aspect of the environment may have impacted results. For example, caregiver-1 said: “*I would attribute some of the alertness… to the rigid schedule [that] does keep him at his best…the constant stimulation of people is different than at home*.*”* Similarly, caregiver-7 noted: “*Here the chair styles are a little bit different, a little deeper and the floors are a little slicker, footwear was a little different*.”

Subject 8 responded dramatically to levodopa but not PF-2562 (see Tables 2-3). Prior to unblinding, both the clinician and caregiver felt Test Period 2 (levodopa) was far superior to Test Period 1 (PF-2562). After discussion with the research ethics consult service, it was decided it was our responsibility to convey these results to the family. This was done and the patient’s family decided to restart levodopa. They reported levodopa had no beneficial effect and the patient remained in a “deep sleep” state.

### Mixed methods results

Integration of quantitative and qualitative data suggested a convergent finding that caregivers favored PF-2562 in four of five patients who completed the study (Table 4). Caregiver observations suggested that alertness and engagement/cognition domains had the most dramatic changes in the four typical LsPD participants. Caregivers also noted that environmental factors likely played a role in the improvements during both weeks. Additionally, the qualitative data uncovered a potential side effect that was not measured discretely in questionnaires (‘twitching’) or detected on quantitative measures. This observation was reported during both the levodopa and PF-2562 testing periods. No caregivers or patients commented specifically on dyskinesia or a special “feeling” that would suggest they were taking levodopa.

**Table 4.**
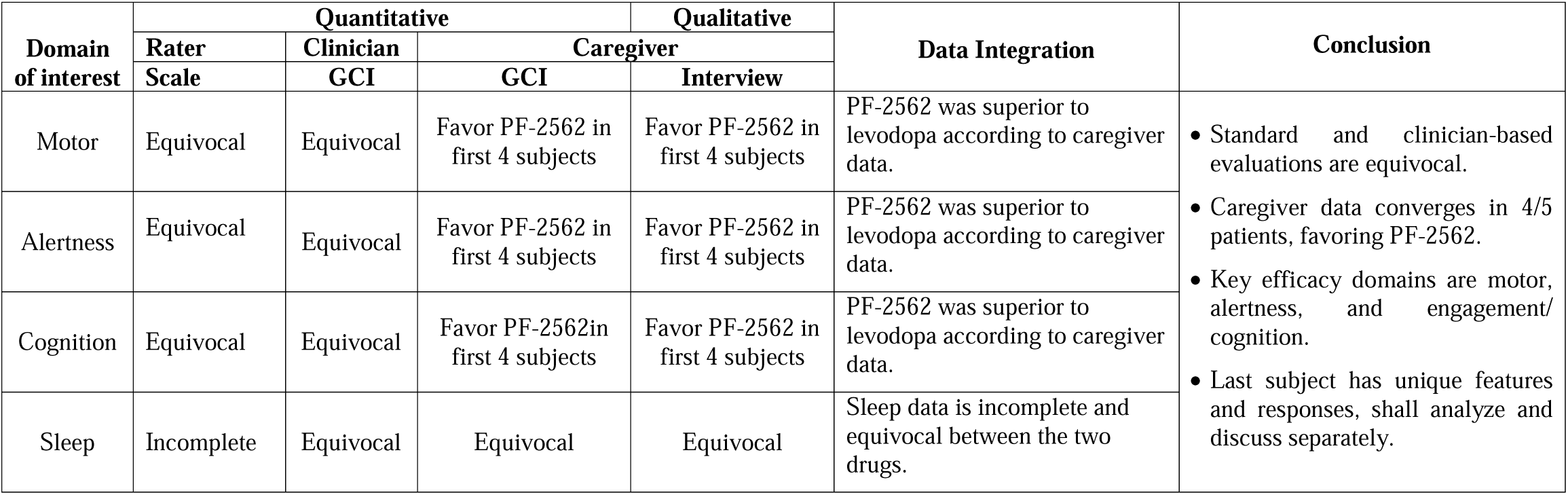
Mixed methods integrated joint display merging quantitative and qualitative data and conclusions.

Overall, caregivers were consistent in their quantitative observations with minimal variability, whereas clinician impressions displayed substantial variability and diverged from caregiver impressions in two of five patients. The rater-dependent standard metrics detected no differences and were not contributory to the overall results.

## Discussion

LsPD patients have many unmet needs, and supportive and palliative care increasingly have been recognized as the best option (e.g., reviews^47,48^). This first controlled interventional study in LsPD patients explored if a D_1_ agonist may have potential benefits in this population exceeding those of palliative care.^24^ We included caregiver perspectives and used mixed methods^49,50^ to identify potential efficacy domains of PF-2562 based on the premise that: 1) traditional clinical tools would be relatively insensitive given the small number of patients and short duration/evaluation period,^38^ and 2) PD patients and their neurologists differ markedly in assessing physical, psychological, and other domains that predict quality of life (QoL).^51^ Our data showed caregivers captured potential benefits of PF-2562 in LsPD patients in ways standard clinical metrics did not. Moreover, mixed methods allowed transformation of semi-structured caregiver observations to quantifiable metrics and identified key domains of improvement (motor, alertness, and cognitive engagement) that warrant future attention.

### The promise of D_1_ agonists in LsPD

Lack of effective treatments for LsPD patients has been a major motivation for seeking disease-modifying therapies^52^ whose clinical trials have focused on enrolling newly-diagnosed or early PD patients. LsPD patients (∼5% of the PD population) have major unmet medical needs and are excluded from most clinical trials by both design and their overall health status. In NHP models^23^ and PD itself,^25^ however, selective D_1_-like agonists are at least equally effective to levodopa.^53^ Importantly, the full D_1_ agonist dihydrexidine markedly attenuated parkinsonian motor signs in MPTP-treated NHPs with disability analogous to subjects in the current study, whereas neither levodopa, nor the D_2/3_ agonist bromocriptine, was effective.^24^

The availability of newer generation D_1/5_ agonists for clinical trials renewed a broad interest in targeting D_1_-like receptors to improve cognitive function in multiple disease states (see review^54^). The current data provide tantalizing evidence that this benefit may extend to LsPD patients. As with motor signs, increased apathy as PD progresses also is observed commonly.^55^ PD apathy and impulse control disorders may be opposite motivational expressions caused by hypo- and hyperdopaminergia, with apathy resulting from hypodopaminergia along with anhedonia, anxiety, and depression. Since the approved D_2_/D_3_ agonists are relatively ineffective, the current data suggest D_1_ agonists also may be effective for motivational deficits in LsPD.

The unique background and response of subject 8 were profound and possibly informative. Since this individual had been largely unresponsive to all treatment in prior years, the dramatic improvement during the levodopa week could be random. It is possible, however, the participant had somehow re-sensitized to levodopa after a three year “drug holiday.” This seems unlikely since there was no effect when the family resumed levodopa. Although highly speculative, another hypothesis is that the two-day period of PF-2562 administration “primed” dopamine circuitry (e.g., by improving sleep structure) to respond more normally to even small amounts of dopamine from a subsequent levodopa challenge six days later. Coupled with the very consistent beneficial responses of the other four patients, the hypothesis is a high priority for further testing, as there are tens-of-thousands of such LsPD patients in the US alone.

### The importance of caregiver perspectives

Most clinical trials rely upon informed clinician judgement based on validated instruments and (when available) imaging/molecular/biochemical markers, but no validated standard scales exist for LsPD.^38^ Clinical ratings of complex behaviors necessarily involve short evaluation epochs with inherent inter-individual and inter-location variability. Prizer et al.^51^ found PD patients and their neurologists differed markedly in assessing physical, psychological, and other domains predicting QoL, and the value of caregiver input has been recognized previously.^33-36,51,56^ In our study, alertness, social interaction, and QoL improvements reported by caregivers reflect changes that are critical to palliative care in LsPD. Decreasing apathy, increasing arousal, or similar improvement in non-motor and motor function could have wide applicability in the absence of “cures.”

In our study, caregiver observations also were more consistent and less variable than experienced physicians. This is not surprising since caregivers were intimately familiar with nuanced baseline patient behaviors, were able to provide insight and context for typical or atypical behavioral observations, and were at the bedside 24/7 during the study. It also is noteworthy that blinded caregivers consistently identified the levodopa week as not being remarkably different from home baseline. This gives credence to the caregivers’ observations and objectivity.

### Utility of mixed methods in a phase I study

Mixed method approaches in clinical trials often are limited to pre-trial use or assessing implementation issues such as recruitment,^57^ and seldom have been used to examine drug trial outcomes.^58^ Our approach revealed efficacy endpoints and observations that were not captured by questionnaires with pre-specified areas of inquiry or anticipated prior to study initiation. The qualitative data added texture to quantitative caregiver evaluations, and their convergence provides compelling data for additional studies investigating PF-2562 to enhance both motor function and cognitive engagement. Future studies should consider integrating mixed method strategies at the phase I stage that may lead both to cost savings and more effective selection of efficacy endpoints in phase II-III trials.

### Limitations and future directions

We are cognizant of the limitations of the current report, but these should be viewed in context. With no prior experience available for interventional studies in LsPD, the design^32^ had to focus on the safety and feasibility, thus limiting both the number of subjects and permitted treatment duration (two days). Efficacy was a second primary endpoint, but it could be evaluated only if the *a priori* safety concerns had not terminated the study. Moreover, the study was completely blinded, all data were locked, and the analysis plan decided *a priori*. The finding of an average significant improvement required excluding the data from subject 8. We feel this was justified as described earlier. It is noteworthy that for the other four subjects, PF-2562 could have worsened them or had no effect, yet these four subjects all had meaningfully better response to PF-2562 than levodopa. We feel that these data provide compelling evidence for further investigation into the potential value of a D_1/5_ agonist in LsPD using increased numbers of subjects and longer drug administration periods. Such studies must incorporate caregiver perspectives that should be conducted at home to eliminate environmental influences on patient behavior. Finally, the few currently available D_1/5_ agonists differ in pharmacological properties (i.e., both intrinsic activity at canonical pathways or functional selectivity).^59,60^ As experience with D_1/5_ agonists in this population is gained, there may be ways to choose compounds with specific profiles for maximal therapeutic benefit.^61-63^

## Data Availability

De-identified data that support the findings of this study are available upon reasonable request from the corresponding author (XH). The data are not publicly available due to privacy or ethical restrictions. All requests must be in writing and will be evaluated in a timely manner by the TBRC executive team.

## Acknowledgements

We express deep gratitude to all of the patients and their caregivers for participating in the study. The completion of the study was made possible by an exceptional team including Shirlynn Mottilla, Nancy Handley, Brenda Hershey-Fell, Lyndsey Houser, Carrie Criley, Patricia Cain, Sue Kocher, Lisa Zeno, Alison Enimpah, Kelly Hoffman, Lori Martin, Brynn Vanderveer, Dina Angello, Kiley Klock, Fiona Fortman, Sandra Franz, Crystal DeMedici, Sarah Debold, and Natalya Knapp. We thank Margaret Hopkins and Dr. K. Randy Young for assisting with the qualitative and mixed methods analysis; Dr. Andrew Foy for consulting on cardiology and ECG questions raised during screening and the conduct of the project; Sara Marlin for serving as our data monitor; and Dr. Gary Thomas for serving on the Data Safety Board (DSB). In addition, we thank Dr. Jennifer McCormick, research ethicist, for providing critical input on our decision to unblind participant 8. This work was supported by a grant from Pfizer Central Research who also supplied test drug and placebo. This work also was supported in part by the National Institute of Neurological Disorders and Stroke and Parkinson’s Disease Biomarker Program (NS060722, NS082151 to XH), and the Penn State National Center for Advancing Translational Sciences, National Institute of Health, through the grant UL1 TR002014. The authors also thank the Quantitative Mixed Methods Center of the Penn State College of Medicine for their assistance with the mixed methods analysis. All analyses, interpretations, and conclusions are those of the authors and not the research sponsors.

## Author Contributions

The trial design was initiated by the authors from Penn State and finalized in collaboration with the Pfizer authors as listed below.

1. Research project: A: Conception, B: Organization, C: Execution
2. Data and Statistical analysis: A: Design, B: Execution, C: Review and critique
3. Manuscript preparation: A: Writing the first draft, B: Review and critique Mechelle M. Lewis: 1A, 1B, 1C, 2A, 2C, 3A, 3B

Lauren Jodi Van Scoy: 1A, 1B, 1C, 2A, 2B, 2C, 3A, 3B

Sol De Jesus:1C, 2C, 3B

Jonathan Hakun: 1B, 1C, 3B

Lan Kong: 2A, 2B, 2C, 3B

Yang Yang: 2C, 3B

Bethany Snyder: 1B, 1C, 2B, 2C, 3B

Sridhar Duvvuri: 2A, 2C

David Gray: 1A, 2A, 2C, 3B

Richard Mailman, 1A, 1B, 2C, 3A, 3B

Xuemei Huang: 1A, 1B, 1C, 2A, 2C, 3A, 3B.

## Use of vertebrate animals or higher non-vertebrate species

This study was done in human Parkinson’s disease patients. No laboratory animals were used.

## Supplemental Information

### Supplemental Text A: Details of Quantitative Analysis

#### Standard scales

As noted in the text, the standardized validated scales assessing alertness, cognition, motor function, and sleep did not detect a significant pattern of differences between levodopa and PF-2562. The clinician ratings were also equivocal (Table 2). <u>For alertness,</u> levodopa improved GCS scores in two participants (1 and 4), worsened them in one (8), and had no effect in two (3 and 7), whereas PF-2562 worsened scores in three participants (1, 4, and 8) and had no effect in two (3 and 7). Levodopa improved SSS scores in 4 participants (1, 4, 7, and 8) and had no effect in one (3), whereas PF-2562 improved SSS score in one participant (8), worsened two (1 and 4), and had no effect in two (3 and 7). <u>For cognition,</u> levodopa worsened SIB scores for two participants (3 and 4) and had no effect in three (1, 7, and 8), whereas PF-2562 improved SIB scores in two participants (3 and 4) and had no effect in three (1, 7 and 8). Levodopa improved FAB scores in three participants (3, 4, and 7) and worsened them in two (1 and 8), whereas PF-2562 worsened FAB scores in three participants (3, 4, and 7) and had no effect in two (1 and 8). <u>For motor function,</u> levodopa improved MDS-UPDRS-III scores in two participants (3 and 4) and worsened them in three (1, 7, and 8), whereas PF-2562 improved MDS-UPDRS-III scores in two participants (1 and 3) and worsened them in three (4, 7, and 8). <u>For sleep efficiency,</u> levodopa improved scores in two participants (7 and 8) and worsened them in one (1), whereas PF-2562 improved sleep efficiency in one participant (7) and worsened it in two (1 and 8).

#### Clinician CGI Ratings

For alertness, clinician GCI-C scores favored levodopa in one participant (8), PF-2562 in two (3 and 4), and were equivocal in two (1 and 7; Table 2). Clinician GCI-C scores for cognition favored levodopa in one participant (8), PF-2562 in two (3 and 4), and were equivocal in two (1 and 7). For motor function, GCI-C scores favored levodopa in one participant (1), PF-2562 in three (3, 4 and 8) and were equivocal in one (7). Clinician GCI-C scores for sleep efficiency favored levodopa in two participants (1 and 8) and were equivocal in three (3, 4, and 7).

#### Caregiver CGI Ratings

For alertness, caregiver GCI-C scores favored levodopa in one participant (8) and PF-2562 in four (1, 3, 4, and 7; Table 2). Caregiver GCI-C scores for cognition favored levodopa in one participant (8) and PF-2562 in four (1, 3, 4, and 7). For motor function, caregiver GCI-C scores favored levodopa in two participants (1 and 8) and PF-2562 in three (3, 4, and 7). Caregiver GCI-C scores for sleep efficiency favored levodopa in one participant (8) and PF-2562 in four (1, 3, 4, and 7).

### Supplemental Text B: Details of Qualitative Analysis

As noted in the text, a conventional content analysis approach that included data transformation was used to evaluate the data.^44^ Published guidelines for methodological rigor of qualitative analysis were followed to ensure attention to the truth-value, applicability, consistency, and neutrality of the findings ^44,46^. Three independent, blinded analysts used qualitative software (NVivo Ver. 11.0, QSR International, Melbourne Australia) to code and analyze the data. **First**, a preliminary codebook was developed inductively based on the common concepts that emerged from the data. The codebook followed closely the structured interview domains yet also included unexpected categories and concepts that were included in the final codebook. **Second**, the preliminary codebook was applied to an additional three transcripts and minor codebook adjustments were made to fit the additional data. Some domains were collapsed as appropriate based on the data. Data saturation (the point at which no new codes emerge) was achieved after reviewing 6 of 12 transcripts (one per participant per treatment week) and the final codebook contained codes for each of the key efficacy domains as well as caregiver observations from the home and study environments. **Third**, the finalized codebook then was used to recode the entire dataset by two coders. Codes were adjudicated by a third analyst to ensure inter-rater reliability. Discrepancies were reconciled via group discussions. **Finally**, analysts used data transformation to convert the qualitative data into categories (i.e., improved, worsened, remained unchanged) for each domain based on the codebook. Any differences in coding was reconciled by group discussion ^64^.

**Supplemental Figure A.**
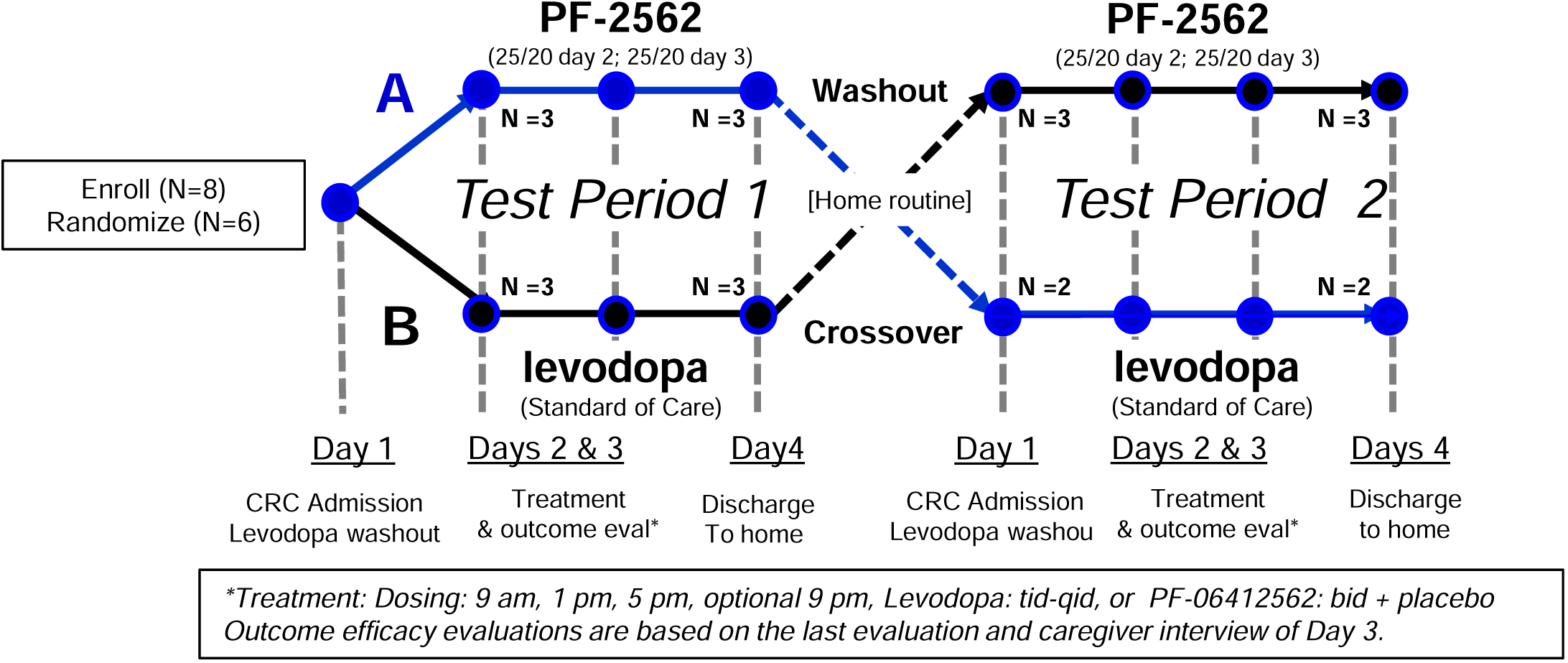
Study schematic. This is a schematic of the overall design of the study (modified from ^32^). Subjects were randomized to receive PF-2562 followed by levodopa (Sequence A, top) or levodopa followed by PF-2562 (Sequence B). PF-2562 (5 mg) or placebo tablets were provided by Pfizer. Sinemet (carbidopa/levodopa, 25/100 mg) tablets were encapsulated to preserve study blind. The bottom part of the schematic shows the events that occurred on each day. Levodopa dose was based on home dosage and regimen. Some subjects received a fourth dose of levodopa if that was required according to their pretrial dosing regimen. Outcome efficacy data are based on the last evaluation and caregiver interview of Day 3.

**Supplemental Table A.**
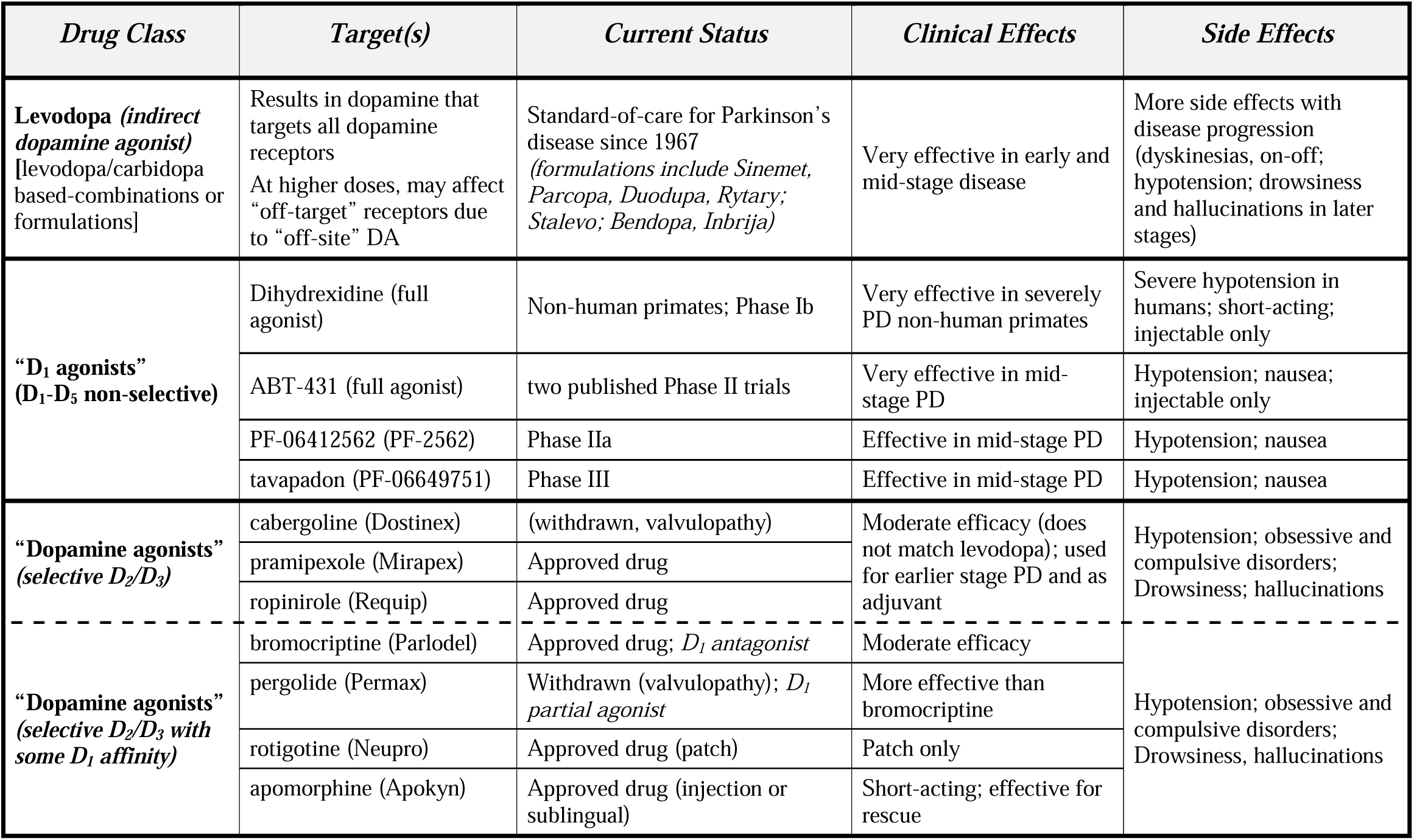
Direct or indirect dopamine receptor agonists that have been approved or in PD clinical trials.

**Supplemental Table B.**
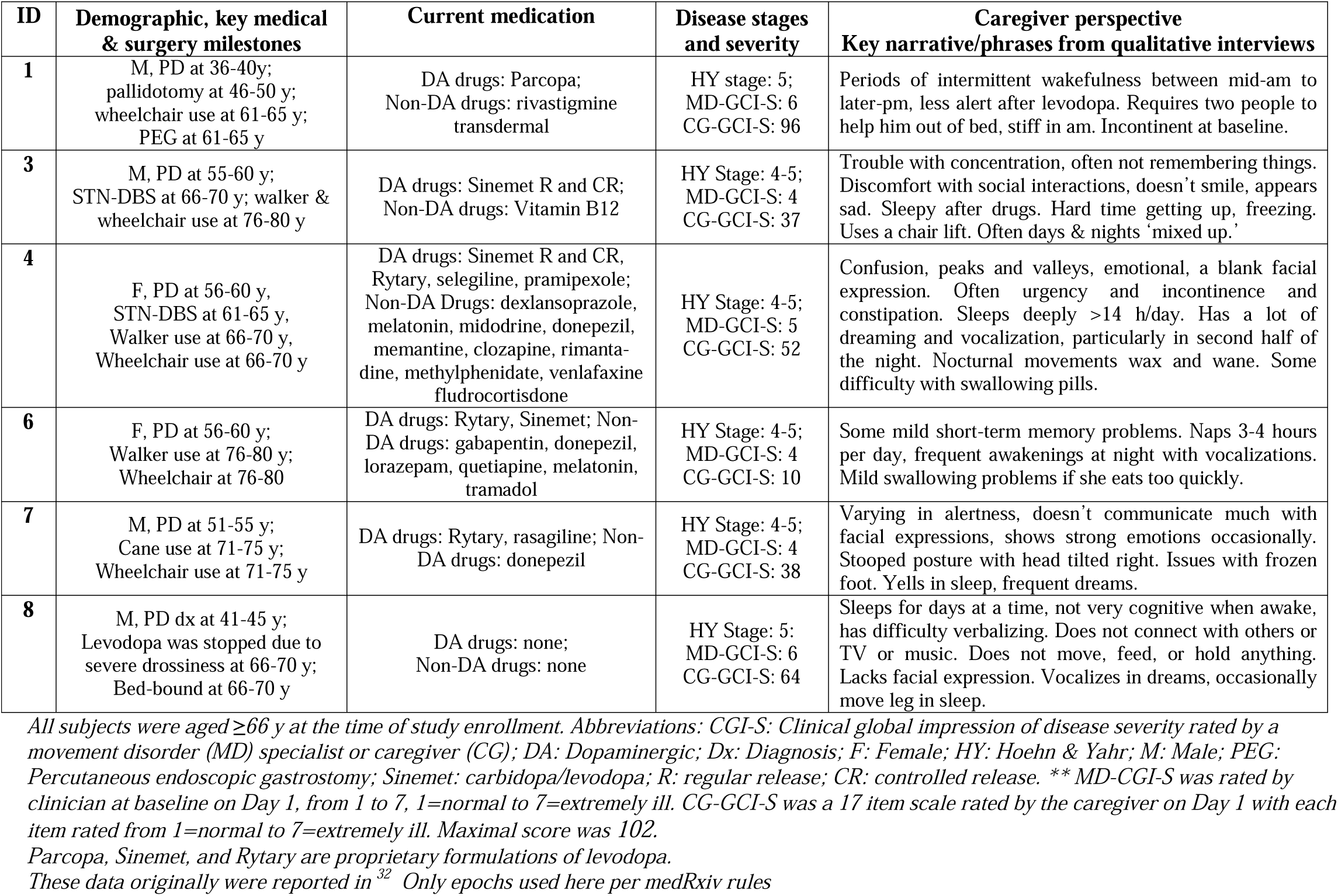
Demographic, clinical history, and baseline data for the randomized participants.

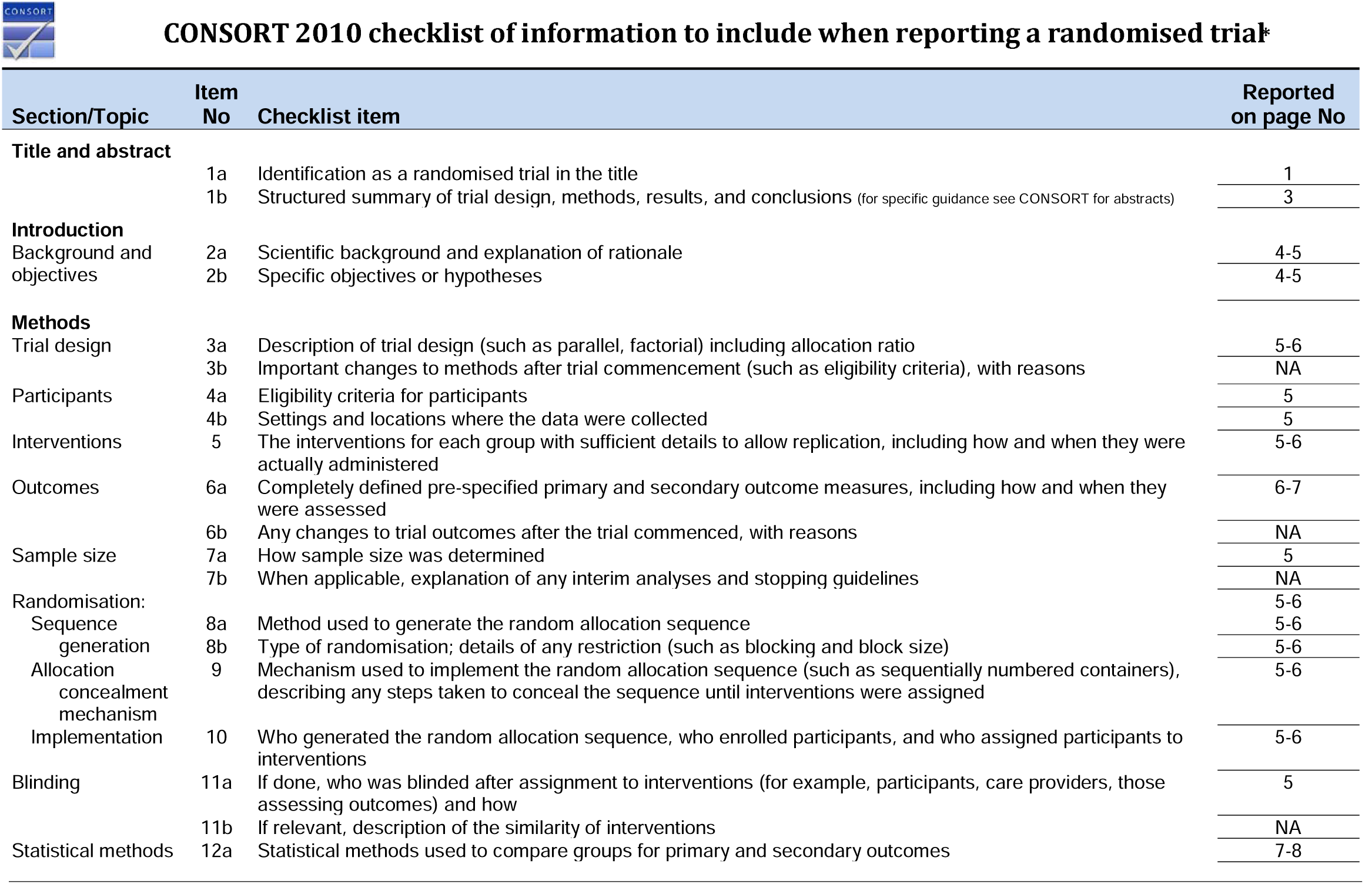
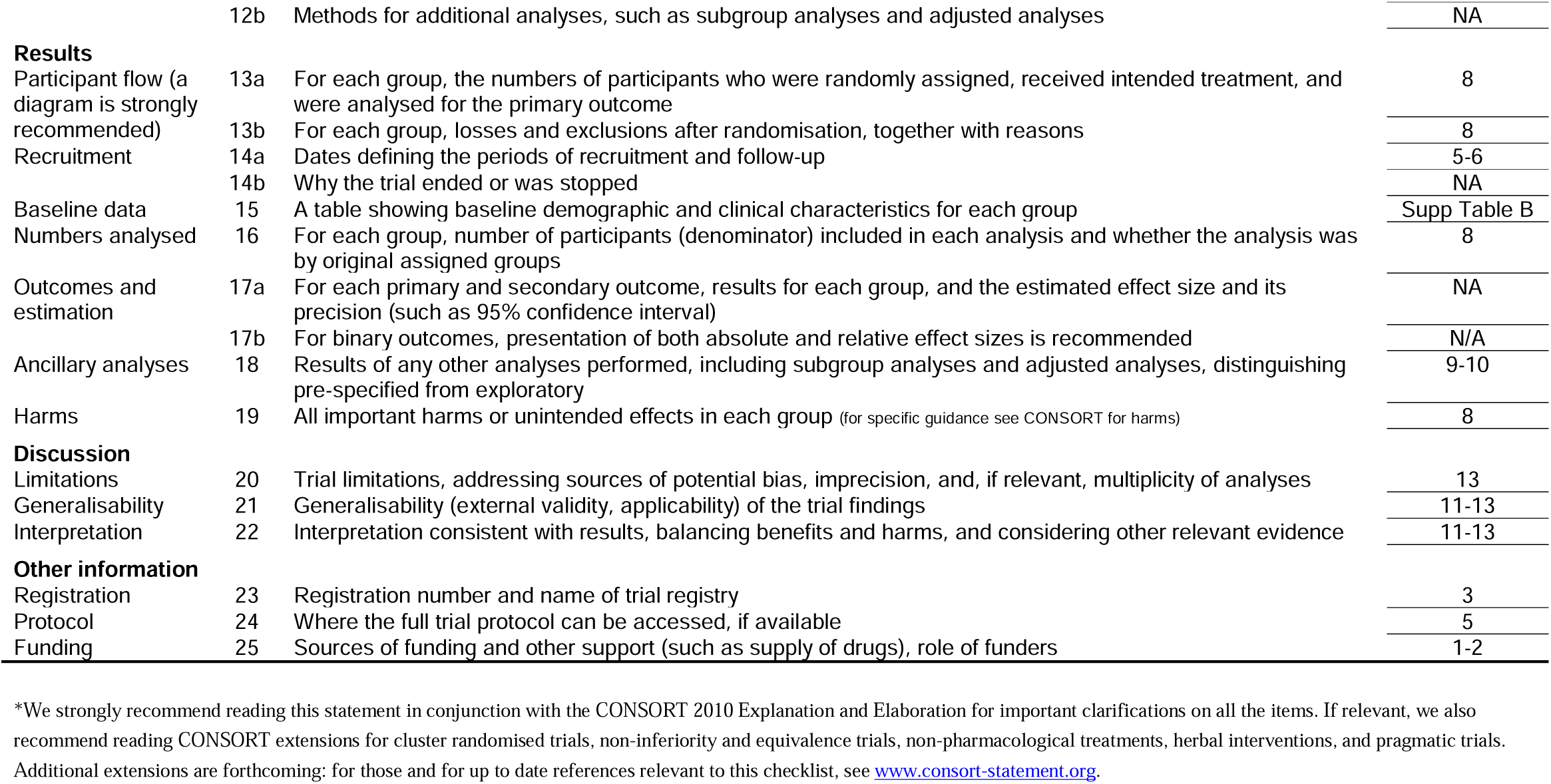

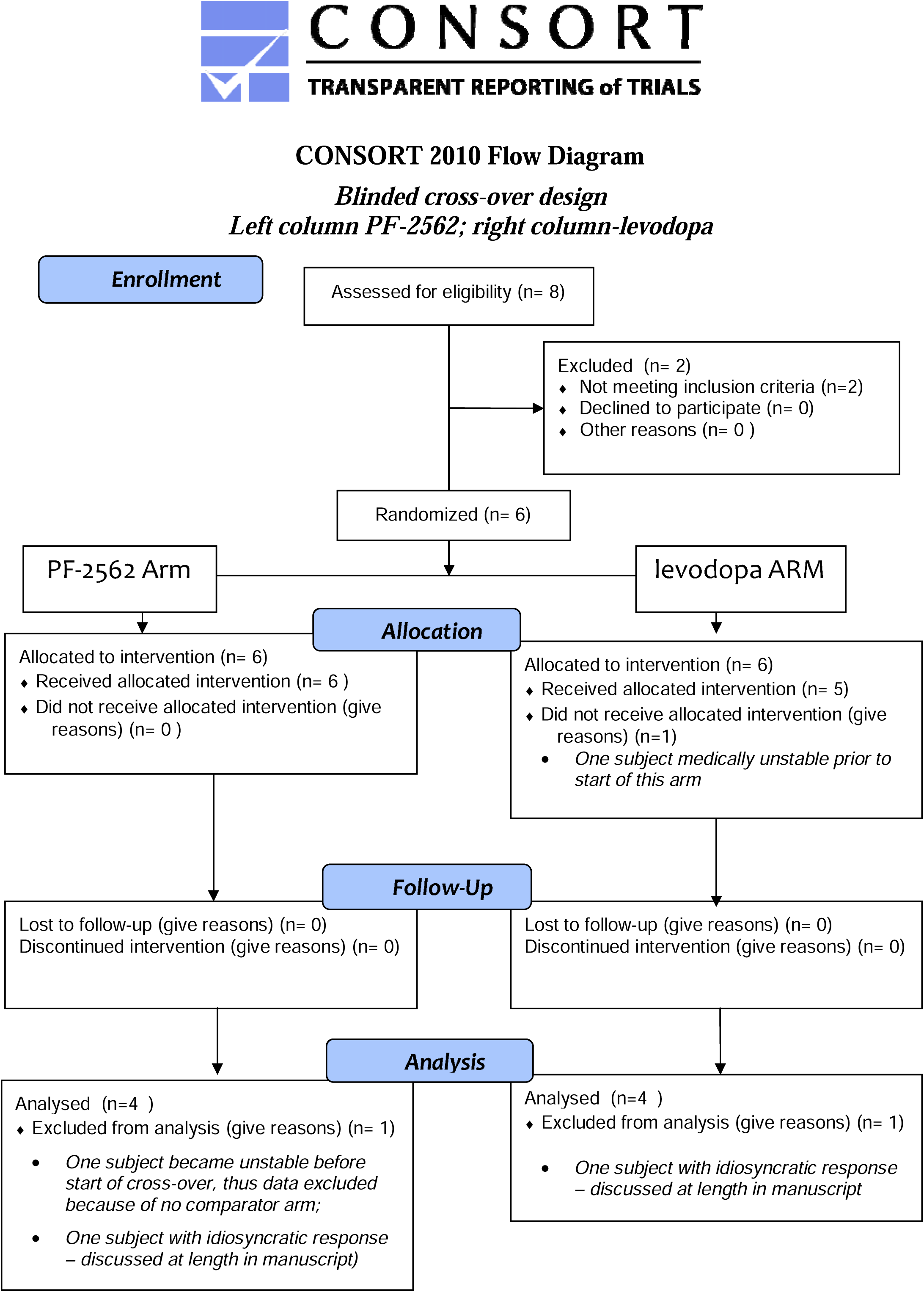

